# Preserved T cell responses to SARS-CoV-2 in anti-CD20 treated multiple sclerosis

**DOI:** 10.1101/2021.10.11.21264694

**Authors:** Tatjana Schwarz, Carolin Otto, Terry C. Jones, Florence Pache, Patrick Schindler, Moritz Niederschweiberer, Felix A. Schmidt, Christian Drosten, Victor M. Corman, Klemens Ruprecht

## Abstract

**Objective:** To analyze humoral and cellular immune responses to SARS-CoV-2 vaccinations and infections in anti-CD20 treated patients with multiple sclerosis (pwMS).

**Methods:** 181 pwMS on anti-CD20 therapy and 41 pwMS who began anti-CD20 therapy were included in a prospective, observational, single-center cohort study between March 2020 and August 2021. 51 pwMS under anti-CD20 treatment, 14 anti-CD20 therapy-naïve pwMS and 19 healthy controls (HC) were vaccinated twice against SARS-CoV-2. We measured SARS-CoV-2 spike protein (full-length, S1 domain and receptor binding domain) immunoglobulin (Ig)G and S1 IgA and virus neutralizing capacity and avidity of SARS-CoV-2 antibodies. SARS-CoV-2 specific T cells were determined by interferon-γ release assays.

**Results:** Following two SARS-CoV-2 vaccinations, levels of IgG and IgA antibodies to SARS-CoV-2 spike protein as well as neutralizing capacity and avidity of SARS-CoV-2 IgG were lower in anti-CD20 treated pwMS than in anti-CD20 therapy-naïve pwMS and in HC (*p*<0.003 for all pairwise comparisons). However, in all anti-CD20 treated pwMS vaccinated twice (n=26) or infected with SARS-CoV-2 (n=2), in whom SARS-CoV-2 specific T cells could be measured, SARS-CoV-2 specific T cells were detectable, at levels similar to those of twice-vaccinated anti-CD20 therapy-naïve pwMS (n=7) and HC (n=19). SARS-CoV-2 S1 IgG levels (*r*=0.42, *p*=0.002), antibody avidity (*r*=0.7, *p*<0.001) and neutralizing capacity (*r*=0.44, *p*=0.03) increased with time between anti-CD20 infusion and second vaccination. Based on detection of SARS-CoV-2 antibodies, SARS-CoV-2 infections occurred in 4/175 (2.3%) anti-CD20 treated pwMS, all of whom recovered fully.

**Interpretation:** These findings should inform treatment decisions and SARS-CoV-2 vaccination management in pwMS.

## Introduction

Management of immunotherapies and vaccinations against severe acute respiratory syndrome coronavirus-2 (SARS-CoV-2) have become key issues in the clinical care of patients with multiple sclerosis (pwMS) during the current SARS-CoV-2 pandemic.^1,2^ Of particular concern, B-cell depleting anti-CD20 therapies have been associated with an increased risk of infections and decreased humoral immune responses to vaccinations.^1–4^ Indeed, serum levels of SARS-CoV-2 immunoglobulin (Ig)G following SARS-CoV-2 vaccinations and infections are reduced in anti-CD20 treated pwMS.^5–11^ Nevertheless, in addition to antibody levels, protection from infection, and especially from severe courses of coronavirus disease-19 (COVID-19), may depend on neutralizing capacity and avidity of anti-SARS-CoV-2 antibodies as well as on induction of SARS-CoV-2 specific T cells.^12–15^ However, data on the neutralizing capacity and avidity of anti-SARS-CoV-2 antibodies in anti-CD20 treated pwMS are scarce.^10^ While very recent studies carried out contemporaneously to this work suggest that anti-CD20 treated pwMS may develop SARS-CoV-2 specific T-cellular immune responses, given the importance of these findings, independent confirmation appears warranted.^9,10^

The first laboratory-confirmed patient with COVID-19 in Germany was reported on January 27, 2020.^16^ To monitor SARS-CoV-2 specific humoral immune responses in anti-CD20 treated pwMS, we started to prospectively collect sera for SARS-CoV-2 antibody determinations from pwMS treated with anti-CD20 therapies at the MS outpatient clinic, Charité Campus Mitte, Charité - Universitätsmedizin Berlin, Germany, on March 5, 2020. Following the start of the SARS-CoV-2 vaccination campaign in Germany in January 2021, we also measured T cell responses to SARS-CoV-2 by interferon-γ release assays (IGRA), permitting the assessment of SARS-CoV-2 specific T cells in routine diagnostics.^17^

Here, we report the results of detailed analyses of humoral and cellular immune responses to SARS-CoV-2 vaccinations and infections in a cohort of a total of 222 anti-CD20 treated pwMS followed through the first 17 months of the SARS-CoV-2 pandemic.

## Patients and Methods

### Patients

This prospective, observational, single-center study was approved by the ethical committee of Charité - Universitätsmedizin Berlin (EA2/152/21 and EA1/068/20) and conducted at the multiple sclerosis outpatient clinic, Charité Campus Mitte, Charité - Universitätsmedizin Berlin, Berlin, Germany, between March 5, 2020, and August 6, 2021 (herein referred to as the study period). Inclusion criteria were age ≥17 years, a diagnosis of relapsing-remitting MS (RRMS) or primary progressive MS (PPMS) according to the McDonald 2017 criteria^18^, and treatment with at least one intravenous infusion of anti-CD20 therapy (ocrelizumab or rituximab) as part of routine clinical care during the study period. Anti-CD20 therapy was initiated either before or after the start of the study. In patients in whom anti-CD20 therapy with ocrelizumab had already been initiated before March 5, 2020, it was continued with intravenous infusions of 600 mg ocrelizumab every 6 months. In patients in whom anti-CD20 therapy was initiated during the study period, it was administered with two intravenous infusions of 300 mg ocrelizumab on day 1 and day 14, followed by intravenous infusions of 600 mg ocrelizumab every 6 months. Some patients had been treated with intravenous rituximab before March 5, 2020, and continued to be treated with 1000 mg rituximab intravenously every 6 months during the study period. Some patients had previously been treated with rituximab at some point during the course of their disease and switched to ocrelizumab following the approval of ocrelizumab by the European Medicines Agency in January 2018. In these patients, 600 mg of ocrelizumab were administered intravenously 6 months after the last infusion of rituximab and every 6 months thereafter. In some patients, treatment intervals with ocrelizumab were extended to longer than 6 months on an individual basis.

On the day of anti-CD20 infusions, a medical history was obtained. Furthermore, a serum sample for determination of SARS-CoV-2 antibodies was withdrawn before the start of anti-CD20 infusions, processed according to standard operating procedures, and stored at −20°C before further analyses. A lithium heparin blood sample was likewise taken at each anti-CD20 infusion and before the start of the infusion for measurement of total serum IgG (Labor Berlin GmbH, Berlin, Germany). Blood sampling for anti-SARS-CoV-2 antibody and total serum IgG determinations was performed on the same day, except for two patients in whom samples for anti-SARS-CoV-2 antibody determinations were withdrawn 13 and 19 days before samples for total IgG determinations. After the start of the SARS-CoV-2 vaccination campaign in Germany in January 2021, a lithium heparin blood sample was additionally withdrawn to carry out IGRA from March 15, 2021, to July 27, 2021.

Patients were considered to be “under anti-CD20 therapy” if they had received at least one intravenous infusion of anti-CD20 therapy, that is, at least 300 mg ocrelizumab at least two weeks before blood samples were withdrawn. Patients were considered as “before anti-CD20 therapy” if they had received no anti-CD20 therapy before blood samples were withdrawn, but later received at least one infusion of anti-CD20 therapy. Some patients had previously been treated with anti-CD20 therapies followed by an anti-CD20 treatment free interval of >18 months before anti-CD20 therapy was re-initiated during the course of this study. These patients were considered to be anti-CD20 therapy naïve and grouped with the “before anti-CD20 therapy” patients. The patients’ clinical and paraclinical data were retrieved from their medical records. All non-vaccinated SARS-CoV-2 antibody positive pwMS were contacted by phone and asked about symptoms of COVID-19.

Blood samples of hospital employees (HE), recruited among staff of the Department of Virology, Charité -Universitätsmedizin Berlin, were obtained 4-6 weeks after the second vaccination with a COVID-19 vaccine.^17^ Blood samples of HE were processed exactly as those of pwMS.

### Detection of anti-SARS-CoV-2 antibodies

IgG and IgA antibodies specific to the S1 domain of the SARS-CoV-2 spike protein were measured by commercially available anti-SARS-CoV-2 S1 IgG and anti-SARS-CoV-2 S1 IgA ELISAs according to the manufacturer’s instructions (Euroimmun, Lübeck, Germany). All ELISA steps were done by the fully automated EUROIMMUN Analyzer I. Briefly, serum was diluted 1:101 with sample buffer and incubated on spike S1 domain pre-coated microplates for 60 min at 37°C. After washing three times, conjugate solution was pipetted and the plate was incubated for 30 min at 37°C. Following a washing step, substrate solution was incubated in the dark for 30 min at room temperature. To stop the reaction, a stop solution was pipetted into each well and the optical density (OD) was detected at 450 nm. OD ratios were calculated by dividing the detected OD by that of the calibrator in the kit. IgG or IgA OD ratios >1.1 were considered reactive.

To confirm SARS-CoV-2 S1 IgG and IgA ELISA results, ELISA reactive sera were also tested by a recombinant SARS-CoV-2 spike protein-based immunofluorescence test (IFT) as previously described.^19^ Briefly, serum samples were diluted 1:10, 1:100, 1:1,000, and 1:10,000 and incubated on Vero B4 cells expressing the spike protein of SARS-CoV-2. IFT titers ≥10 were considered reactive. Serum from one twice-vaccinated anti-CD20 pwMS was excluded from the IFT analysis due to nonspecific fluorescence.

All samples from vaccinated and infected pwMS and from vaccinated HE were also analyzed by a solid phase immunoassay containing four recombinant SARS-CoV-2 antigens (nucleocapsid, spike receptor binding domain (RBD), S1 domain, and full-length spike; SeraSpot®Anti-SARS-CoV-2 IgG, Seramun Diagnostica, Heidsee, Germany). Measurements were performed according to the manufacturer’s instructions as previously described.^20^

### Neutralizing capacity of anti-SARS-CoV-2 antibodies

To measure the neutralizing capacity of anti-SARS-CoV-2 antibodies, a plaque reduction neutralization test (PRNT) was used as previously described.^19,21^ Briefly, heat-inactivated sera were diluted in OptiPro, mixed 1:1 with 100 plaque forming units of SARS-CoV-2 (strain NCBI GenBank AccNo.: MT270112.01) and incubated on Vero E6 cells seeded in 24-well plates on the previous day. After 60 minutes incubation at 37°C, the supernatant was discarded, cells washed with PBS and overlaid with 1.2% Avicel solution in DMEM. After three days at 37°C, cells were inactivated, fixed with a 6% formaldehyde/PBS solution and stained with crystal violet. Serum dilutions with a plaque reduction of 50% are referred to as PRNT50 titers. PRNT50 titers ≥20 were considered to indicate the presence of neutralizing antibodies. For numerical calculations, PRNT50 titers <20 were set to 10 and >640 to 640.

### Determination of antibody avidity

IgG avidity maturation was detected using a modified SARS-CoV-2 S1 ELISA (Euroimmun) as previously described.^20^ The percentage relative avidity index was calculated as follows: (OD of sample treated with urea/OD of sample treated with PBS) x 100%. Avidity indices between 40-60% were considered as borderline and above 60% as high avidity.

### Interferon-γ release assay (IGRA)

SARS-CoV-2 spike protein specific T cells were monitored utilizing a commercially available IGRA (Euroimmun) according to the manufacturer’s instructions as previously described.^20^ Interferon-γ values higher than the maximum interferon-γ values in the not infected/vaccinated control group (111.41 mIU/ml) were regarded as indicative of the presence of SARS-CoV-2 spike protein specific T cells.

### Total IgG in serum

Total immunoglobulin IgG was determined by immunoturbidimetry at Labor Berlin GmbH, Berlin, Germany. One pwMS with a highly elevated serum IgG (41.5 g/l) due to co-existing multiple myeloma was excluded from further analysis.

### Statistical analyses

Categorical variables are provided as frequencies and percentages with 95% confidence intervals (CI) calculated using the Wilson procedure with a correction for continuity.^22^ Continuous variables are presented as median values and interquartile ranges (IQR). Group comparisons were performed by Chi square tests or nonparametric Mann-Whitney *U* test or Kruskal-Wallis test with Dunn’s multiple comparisons test as appropriate. Correlations of two datasets were assessed by Spearman’s method. *P* values of <0.05 were considered statistically significant. For statistical analyses, GraphPad PRISM (version 9.2.0) was used.

## Results

### Patients and controls

Between March 5, 2020, and August 6, 2021, 222 consecutive pwMS (177 with relapsing-remitting MS [RRMS] and 45 with primary progressive MS [PPMS]) treated with at least one infusion of anti-CD20 therapy (220 with ocrelizumab and 2 with rituximab) at the MS outpatient clinic, Charité Campus Mitte, Charité -Universitätsmedizin Berlin, were included in a prospective observational cohort study. According to treatment status at the date of the latest sample collection, pwMS were grouped into those before initiation of anti-CD20 therapy (n=41) and those under anti-CD20 therapy (n=181). Age, female/male ratio, the proportions of patients with RRMS/PPMS, the expanded disability status scale (EDSS) score, and the percentages of patients with any previous or at least two previous immunotherapies did not differ between pwMS before initiation of anti-CD20 therapy and under anti-CD20 therapy (Table 1). Nineteen hospital employees (HE), who were on average 7.5 years younger than pwMS, were included as controls (Table 1). Figure 1 provides an overview of the pwMS and the blood samples analysed in this work and summarizes the number of pwMS who received one or two SARS-CoV-2 vaccinations as well as the number of SARS-CoV-2 infections among pwMS under and before initiation of anti-CD20 therapy.

**Table 1.**
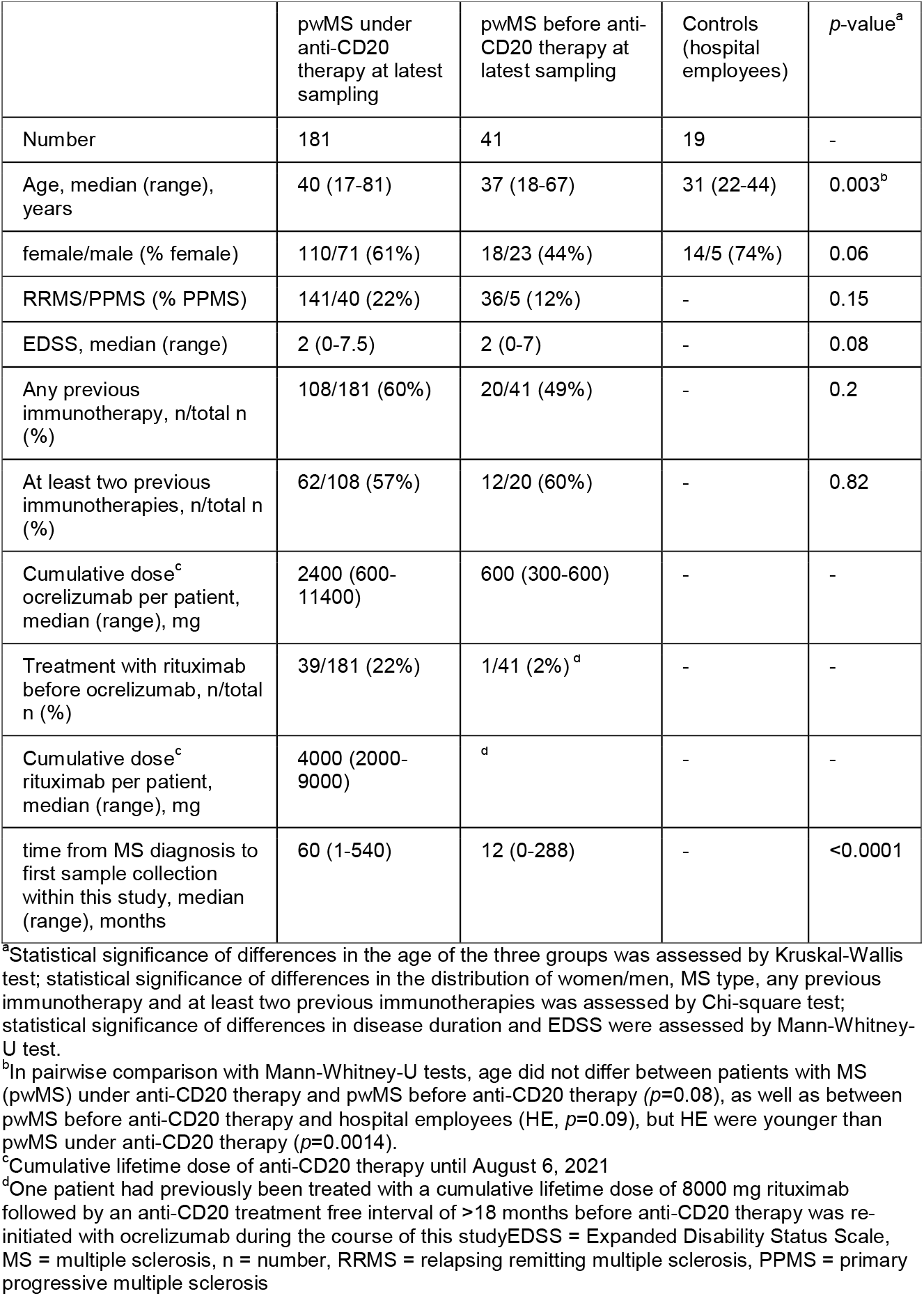
Demographic, clinical and treatment characteristics of patients with multiple sclerosis and demographic characteristics of healthy controls.

|  | pwMS under anti-CD20 therapy at latest sampling | pwMS before anti-CD20 therapy at latest sampling | Controls (hospital employees) | *p*-value<sup>a</sup> |
| --- | --- | --- | --- | --- |
| Number | 181 | 41 | 19 | - |
| Age, median (range), years | 40 (17-81) | 37 (18-67) | 31 (22-44) | 0.003<sup>b</sup> |
| female/male (% female) | 110/71 (61%) | 18/23 (44%) | 14/5 (74%) | 0.06 |
| RRMS/PPMS (% PPMS) | 141/40 (22%) | 36/5 (12%) | - | 0.15 |
| EDSS, median (range) | 2 (0-7.5) | 2 (0-7) | - | 0.08 |
| Any previous immunotherapy, n/total n (%) | 108/181 (60%) | 20/41 (49%) | - | 0.2 |
| At least two previous immunotherapies, n/total n (%) | 62/108 (57%) | 12/20 (60%) | - | 0.82 |
| Cumulative dose<sup>c</sup> ocrelizumab per patient, median (range), mg | 2400 (600-11400) | 600 (300-600) | - | - |
| Treatment with rituximab before ocrelizumab, n/total n (%) | 39/181 (22%) | 1/41 (2%)<sup>d</sup> | - | - |
| Cumulative dose<sup>c</sup> rituximab per patient, median (range), mg | 4000 (2000-9000) | <sup>d</sup> | - | - |
| time from MS diagnosis to first sample collection within this study, median (range), months | 60 (1-540) | 12 (0-288) | - | <0.0001 |

<sup>a</sup>Statistical significance of differences in the age of the three groups was assessed by Kruskal-Wallis test; statistical significance of differences in the distribution of women/men, MS type, any previous immunotherapy and at least two previous immunotherapies was assessed by Chi-square test; statistical significance of differences in disease duration and EDSS were assessed by Mann-Whitney-U test.
<sup>b</sup>In pairwise comparison with Mann-Whitney-U tests, age did not differ between patients with MS (pwMS) under anti-CD20 therapy and pwMS before anti-CD20 therapy ( $p=0.08$ ), as well as between pwMS before anti-CD20 therapy and hospital employees (HE, $p=0.09$ ), but HE were younger than pwMS under anti-CD20 therapy ( $p=0.0014$ ).
<sup>c</sup>Cumulative lifetime dose of anti-CD20 therapy until August 6, 2021
<sup>d</sup>One patient had previously been treated with a cumulative lifetime dose of 8000 mg rituximab followed by an anti-CD20 treatment free interval of >18 months before anti-CD20 therapy was re-initiated with ocrelizumab during the course of this study EDSS = Expanded Disability Status Scale, MS = multiple sclerosis, n = number, RRMS = relapsing remitting multiple sclerosis, PPMS = primary progressive multiple sclerosis

**Figure 1.**
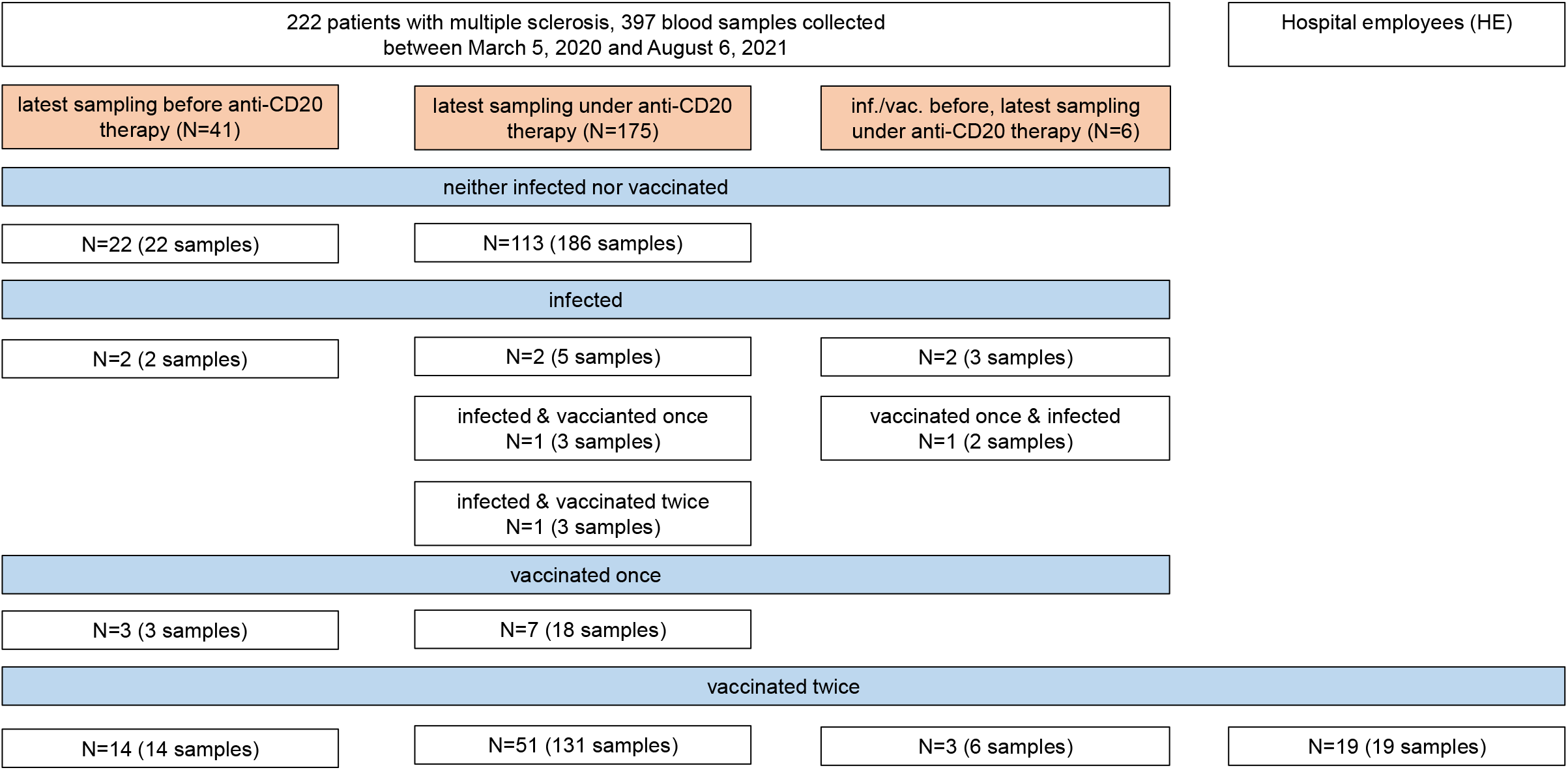
Overview of patients with multiple sclerosis and healthy controls (hospital employees) and blood samples analyzed in this study. Infection with SARS-CoV-2 was defined as detection of reactive SARS-CoV-2 antibodies in at least two of four antibody test systems (anti-SARS-CoV-2 S1 IgG ELISA, anti-SARS-CoV-2 S1 IgA ELISA, spike protein immunofluorescence test, and SeraSpot® anti-SARS-CoV-2 IgG) or a positive interferon-γ release assay (IGRA) in patients with multiple sclerosis not vaccinated against SARS-CoV-2. inf. = infected, vac. = vaccinated

### Humoral immune responses to SARS-CoV-2 vaccinations

Of the 222 pwMS included in this study, a total of 397 serum samples were collected during the study period (Fig 1). Figure 2A shows SARS-CoV-2 S1 IgG reactive samples per study month for all 222 pwMS and Figure 2B shows courses of anti-SARS-CoV-2 S1 IgG levels in individual pwMS. While anti-SARS-CoV-2 S1 IgG reactive samples were only infrequently detected during the peak of the COVID-19 wave in Berlin between October 2020 and January 2021, their frequency increased following the start of the German SARS-CoV-2 vaccination campaign in January 2021.

**Figure 2.**
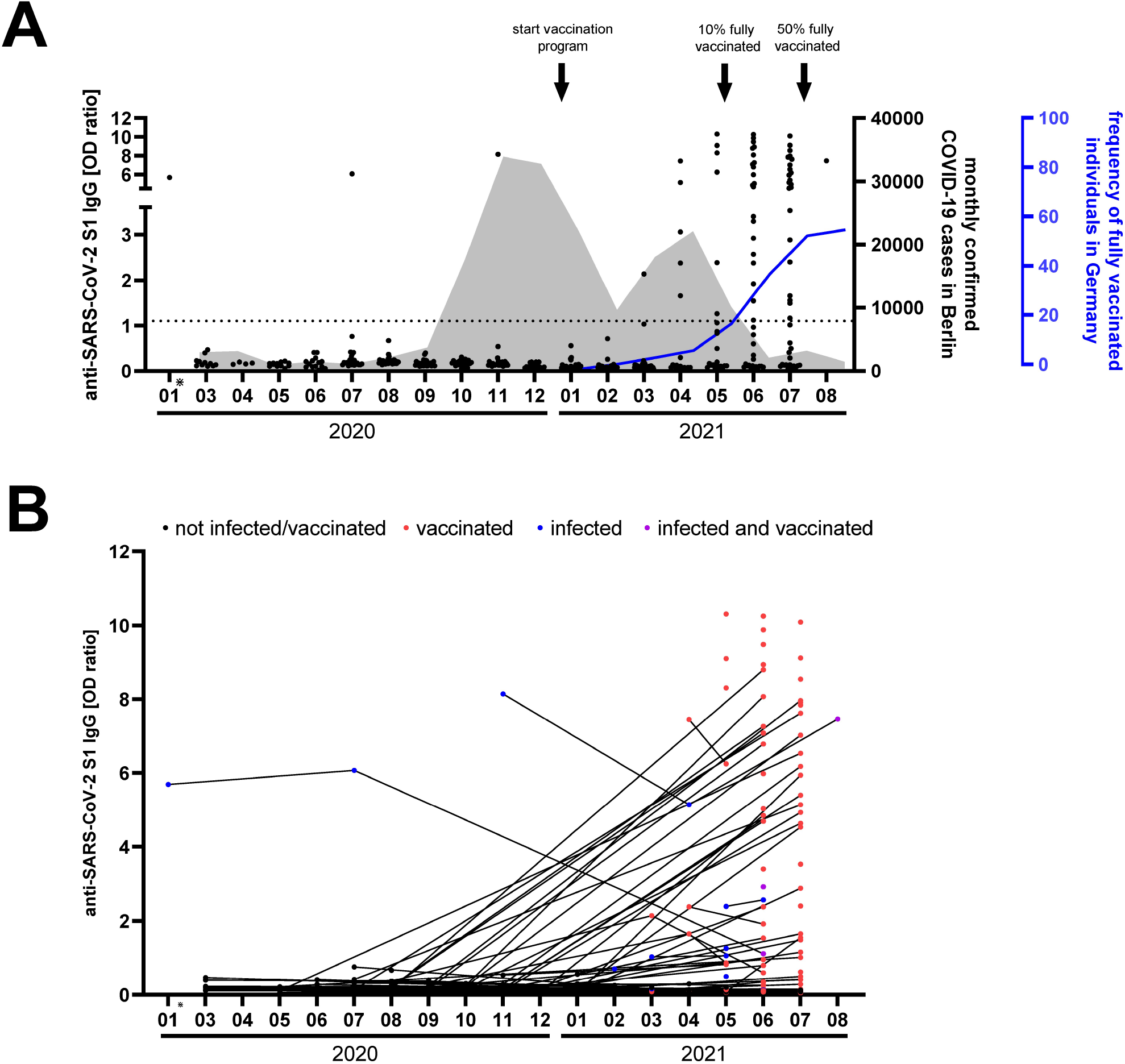
Overview of anti-SARS-CoV-2 S1 IgG responses in patients with multiple sclerosis (pwMS) during the course of the study. Anti-SARS-CoV-2 S1 IgG was measured by a commercial ELISA in 398 serum samples (dots) from 222 pwMS. (A) Levels of anti-SARS-CoV-2 S1 IgG during each month of the study period. The sample from January 2020 is a retrospectively analyzed biobanked sample from a SARS-CoV-2 infected anti-CD20 treated pwMS. The grey area shows monthly confirmed COVID-19 cases in Berlin over the course of the pandemic. The vaccination progress of fully-vaccinated individuals in Germany is depicted as relative frequency (blue line). (B) Individual courses of anti-SARS-CoV-2 IgG S1 levels in 222 pwMS. *biobank sample IgG = immunoglobulin G, OD = optical density, S1 = SARS-CoV-2 spike protein S1 domain

During the study period, 51 pwMS under anti-CD20 therapy and 14 pwMS before initiation of anti-CD20 therapy were vaccinated twice against SARS-CoV-2 (Fig 1). Of the 14 pwMS before initiation of anti-CD20 therapy, 10 were untreated, three treated with dimethyl fumarate and one with glatiramer acetate within the last year. Vaccinations were performed with the BNT162b2 vaccine in 45 of 51 (88.2%, 95%CI: 76.7-94.5) pwMS under anti-CD20 therapy and 10 of 14 (71.4%; 95%CI: 45.4-88.3) pwMS before initiation of anti-CD20 therapy (for further vaccines used, see Figure 4A). For analysis of humoral immune responses to SARS-CoV-2 vaccinations, we used the latest available sample from pwMS (median [IQR] interval from the second vaccination to blood withdrawal: 40 [31-47] days). Nineteen HE vaccinated twice against SARS-CoV-2 with BNT162b2/BNT162b2 (n=7) or ChAdOx1/BNT162b2 (n=12) served as controls (median [IQR] interval from the second vaccination to blood withdrawal: 28 [22-29] days).

After the second SARS-CoV-2 vaccination, the frequency of patients with anti-SARS-CoV-2 S1 IgG antibodies above the assay threshold for reactivity was lower in anti-CD20 treated pwMS (26/51, 50.9%, 95%CI: 37.7-64.3%) than in pwMS before anti-CD20 therapy (13/14, 92.9%, 95%CI: 68.5-97.6%, *p*=0.005) and HE (19/19, 100%, 95% CI: 83.2-100%, *p*<0.0001) (Table 2). Accordingly, anti-SARS-CoV-2 S1 IgG levels were lower in anti-CD20 treated pwMS (median [IQR] OD ratio 1.2 [0.1-5.1]) compared to pwMS before anti-CD20 therapy (9.0 [6.8-9.9], *p*<0.0001) and to HE (8.8 [8.0-9.4], *p*<0.0001) (Fig 3A, Fig 4B, Table 2). No significant difference of anti-SARS-CoV-2 S1 IgG antibodies was detected between pwMS before anti-CD20 therapy and HE (*p*=1). Similar results were obtained for anti-SARS-CoV-2 S1 IgA antibodies (Fig 4C). Likewise, anti-CD20 treated pwMS vaccinated twice had lower IgG levels against a recombinant SARS-CoV-2 spike protein as detected by IFT (Fig 4D). Furthermore, using a microarray based immunoassay, anti-CD20 treated pwMS vaccinated twice had lower IgG levels against the receptor binding domain of the SARS-CoV-2 spike protein and the full-length spike protein (Fig 4E, F). Antibodies against the SARS-CoV-2 nucleocapsid, indicating past SARS-CoV-2 infection, were undetectable in pwMS vaccinated twice (Fig 2G).

**Table 2.** Proportion of positive outcomes in the different test systems.

|  | anti-CD20 therapy |  |  |  |  |  | no anti-CD20 therapy |  |  |  | Infection/vaccination before therapy, sampling after therapy |  |  | HE |
| --- | --- | --- | --- | --- | --- | --- | --- | --- | --- | --- | --- | --- | --- | --- |
| test system | 1<sup>st</sup> vac | 2<sup>nd</sup> vac | infected + 1<sup>st</sup> vac | infected+ 2<sup>nd</sup> vac | infected | Not infected/ vaccinated | 1<sup>st</sup> vac | 2<sup>nd</sup> vac | infected | Not infected/ vaccinated | 2<sup>nd</sup> vac | infected | 1<sup>st</sup> vac + infected | 2<sup>nd</sup> vac |
| IgG S1 ELISA<sup>a</sup> | 0.0 (0.0-35.4) [0/7] | 50.9 (37.7-64.3) [26/51] | 100 (5.1-100) [1/1] | 100 (5.1-100) [1/1] | 50.0 (2.6-97.4) [1/2] | 0.0 (0.0-3.3) [0/113] | 100 (43.9-100) [3/3] | 92.9 (68.5-97.6) [13/14] | 0.0 (0.0-94.9) [0/2] | 0.0 (0.0-14.9) [0/22] | 100 (43.9-100) [3/3] | 50.0 (2.6-97.4) [1/2] | 100 (5.1-100) [1/1] | 100 (83.2-100) [19/19] |
| IgA S1 ELISA<sup>a</sup> | 14.3 (0.7-51.3) [1/7] | 35.3 (23.6-49.0) [18/51] | 100 (5.1-100) [1/1] | 100 (5.1-100) [1/1] | 100 (17.8-100) [2/2] | 3.5 (1.4-8.8) [4/113] | 100 (43.9-100) [3/3] | 85.7 (60.1-97.5) [12/14] | 50.0 (2.6-97.4) [1/2] | 0.0 (0.0-14.9) [0/22] | 66.7 (11.8-98.3) [2/3] | 50.0 (2.6-97.4) [1/2] | 0.0 (0.0-94.9) [0/1] | 94.7 (75.4-99.7) [18/19] |
| IFT titer<sup>b</sup> | 28.6 (5.1-64.1) [2/7] | 54.0 (40.4-67.0) [27/50] | 0.0 (0.0-94.9) [0/1] | 0.0 (0.0-94.9) [0/1] | 100 (17.8-100) [2/2] | 0.0 (0.0-49.0) [0/4] | 100 (43.9-100) [3/3] | 92.9 (68.5-99.6) [13/14] | 100 (17.8-100) [2/2] | n.a. | 100 (43.9-100) [3/3] | 100 (5.1-100) [1/1] | 0.0 (0.0-94.9) [0/1] | 100 (83.2-100) [19/19] |
| RBD spot array<sup>c</sup> | 0.0 (0.0-35.4) [0/7] | 33.3 (22.0-47.0) [17/51] | 100 (5.1-100) [1/1] | 0.0 (0.0-94.9) [0/1] | 50.0 (2.6-97.4) [1/2] | n.a. | 100 (43.9-100) [3/3] | 92.9 (68.5-97.6) [13/14] | 0.0 (0.0-94.9) [0/2] | n.a. | 100 (43.9-100) [3/3] | 50.0 (2.6-97.4) [1/2] | 0.0 (0.0-94.9) [0/1] | 100 (83.2-100) [19/19] |
| spike full spot array<sup>c</sup> | 0.0 (0.0-35.4) [0/7] | 37.3 (25.3-51.0) [19/51] | 100 (5.1-100) [1/1] | 0.0 (0.0-94.9) [0/1] | 50.0 (2.6-97.4) [1/2] | n.a. | 66.7 (11.8-98.3) [2/3] | 92.9 (68.5-97.6) [13/14] | 0.0 (0.0-94.9) [0/2] | n.a. | 66.7 (11.8-98.3) [2/3] | 100 (17.8-100) [2/2] | 0.0 (0.0-94.9) [0/1] | 100 (83.2-100) [19/19] |
| NP spot array<sup>c</sup> | 0.0 (0.0-35.4) [0/7] | 0.0 (0.0-7.0) [0/51] | 100 (5.1-100) [1/1] | 0.0 (0.0-94.9) [0/1] | 0.0 (0.0-82.2) [0/2] | n.a. | 0.0 (0.0-56.2) [0/3] | 0.0 (0.0-21.5) [0/14] | 0.0 (0.0-94.9) [0/2] | n.a. | 0.0 (0.0-56.2) [0/3] | 50.0 (2.6-97.4) [1/2] | 0.0 (0.0-94.9) [0/1] | 0.0 (0.0-16.8) [0/19] |
| PRNT50 titer<sup>d</sup> | n.a. | 69.2 (50.0-83.5) [18/26] | 100 (5.1-100) [1/1] | 0.0 (0.0-94.9) [0/1] | 0.0 (0.0-94.9) [0/1] | n.a. | 100 (43.9-100) [3/3] | 92.3 (66.7-99.6) [12/13] | n.a. | n.a. | 66.7 (11.8-98.3) [2/3] | 100 (5.1-100) [1/1] | 0.0 (0.0-94.9) [0/1] | 100 (83.2-100) [19/19] |
| IgG RAI<sup>e</sup> | n.a. | 15.4 (6.2-33.5) [4/26] | 0.0 (0.0-94.9) [0/1] | 100 (5.1-100) [1/1] | 0.0 (0.0-94.9) [0/1] | n.a. | 0.0 (0.0-56.2) [0/3] | 76.9 (49.7-91.8) [10/13] | n.a. | n.a. | 0.0 (0.0-56.2) [0/3] | 0.0 (0.0-94.9) [0/1] | 0.0 (0.0-94.9) [0/1] | 89.5 (68.6-98.1) [17/19] |
| IGRA S1<sup>f</sup> | 83.3 (43.7 - 99.1) [5/6] | 100 (87.1-100) [26/26] | n.a. | 100 (5.1-100) [1/1] | 100 (17.8-100) [2/2] | 0.0 (0.0-16.1) [0/20] | 100 (43.9-100) [3/3] | 100 (64.6-100) [7/7] | 100 (5.1-100) [1/1] | 0.0 (0.0-27.8) [0/10] | 100 (43.9-100) [3/3] | 100 (17.8-100) [2/2] | 100 (5.1-100) [1/1] | 100 (83.2-100) [19/19] |

Data shown are from the latest available sample during the study period. All values are given as percentage (95% confidence interval) [number reactive samples/total number samples].
<sup>a</sup>IgG or IgA OD ratios >1.1 were considered reactive.
<sup>b</sup>IFT titers ≥10 were considered reactive.
<sup>c</sup>S/Co ratios ≥1.0 were considered reactive.
<sup>d</sup>PRNT50 titers ≥20 were considered to indicate the presence of neutralizing antibodies.
<sup>e</sup>Avidity indices above 60% were used as cut-off.
<sup>f</sup>Interferon-γ values >111.41 mIU/ml were considered positive.
IFN-γ = interferon-γ, IFT = immunofluorescence test, IgA = Immunoglobulin A, IgG = Immunoglobulin G, IGRA = interferon-γ release assay, IU = international units, NP = nucleocapsid protein, RAI = relative avidity index, RBD = receptor-binding domain, PRNT = plaque reduction neutralization test, S1 = spike subdomain 1, S/CO = signal to cut-off, vac = vaccination

**Figure 3.**
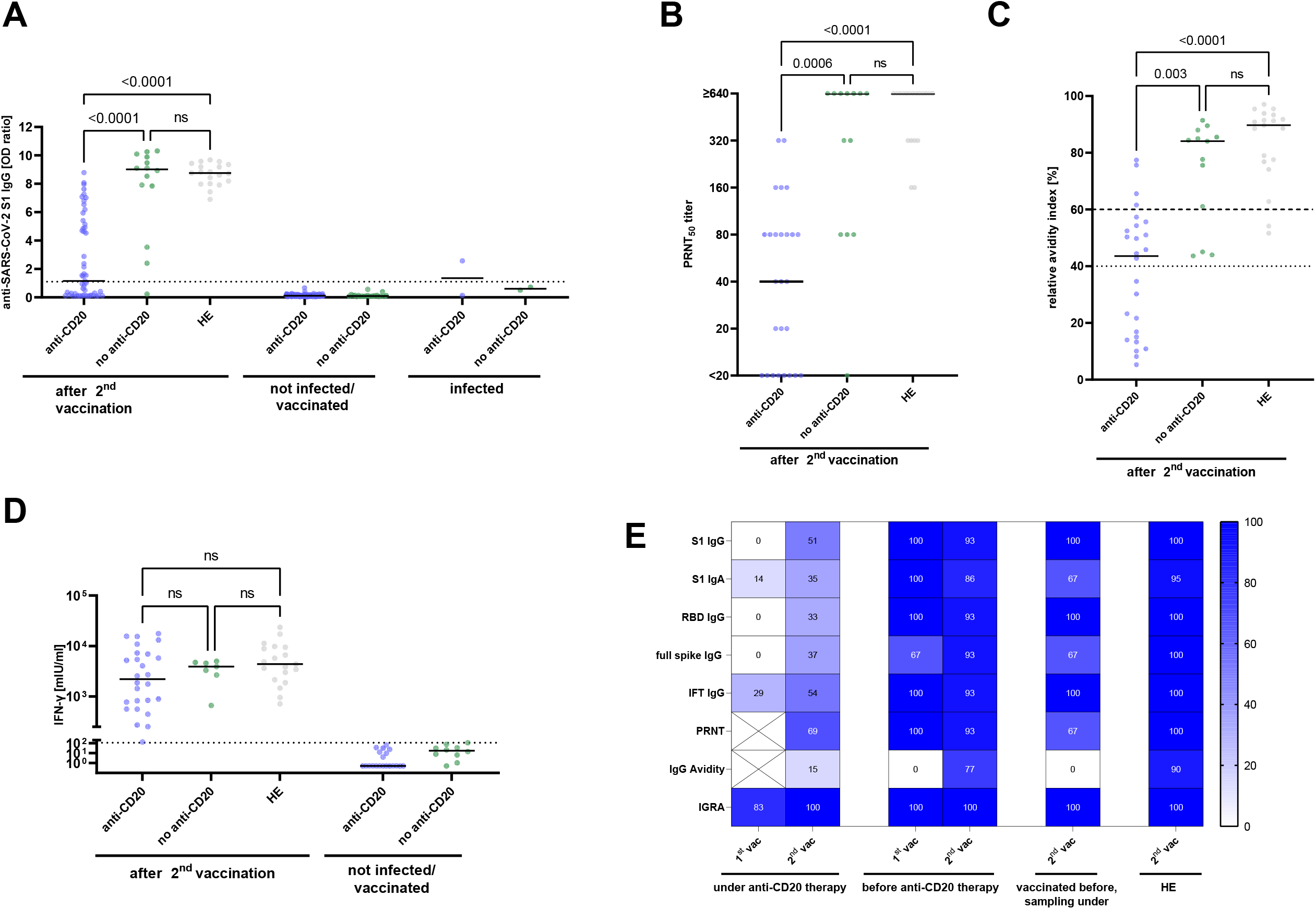
Humoral and cellular SARS-CoV-2 specific immune responses in patients with MS (pwMS) and healthy controls (hospital employees, HE). (A) Sera of anti-CD20 treated pwMS (anti-CD20), pwMS before anti-CD20 therapy (no anti-CD20) and HE were tested for anti-SARS-CoV-2 S1 IgG antibodies. Anti-SARS-CoV-2 specific S1 IgG OD ratios among pwMS after two vaccinations, pwMS who were not vaccinated/infected, and pwMS after SARS-CoV-2 infections are shown. The dotted horizontal line represents an OD ratio of 1.1, levels above which indicate the presence of anti-SARS-CoV-2 S1 IgG. Functionality of SARS-CoV-2 specific antibodies after two vaccinations was analysed in anti-CD20 treated pwMS, pwMS before anti-CD20 therapy, and HE by (B) determination of SARS-CoV-2 neutralizing capacity, using a plaque reduction neutralization test (PRNT), and by (C) IgG avidity maturation. Dotted horizontal lines indicate relative antibody indices >40%, which were considered borderline, and relative antibody indices >60% which were considered high avidity. (D) SARS-CoV-2 specific T cell responses were measured in whole blood samples by IGRA in pwMS and HE vaccinated twice and in pwMS who were not vaccinated/infected. SARS-CoV-2 specific T cells were considered to be present if IFN-γ release was higher than the highest value in the not vaccinated/infected control group (111.41 mIU/ml, dotted horizontal line). (A-D) Horizontal lines indicate the median; *P* values were calculated by the non-parametric Kruskal-Wallis test with Dunn’s multiple comparisons test. (E) Heatmap summarizing the percentage of positive outcomes per test in pwMS and HE (results for groups with n<3 were not included). HE = hospital employees, IFN-γ = interferon-γ, IFT = immunofluorescence test, IgA = immunoglobulin A, IgG = immunoglobulin G, IGRA = interferon-γ release assay, IU = international units, ns = not significant, OD = optical density, PRNT = plaque reduction neutralization test, RBD = receptor binding domain, S1 = SARS-CoV-2 spike protein S1 domain, vac = vaccination

**Figure 4.**
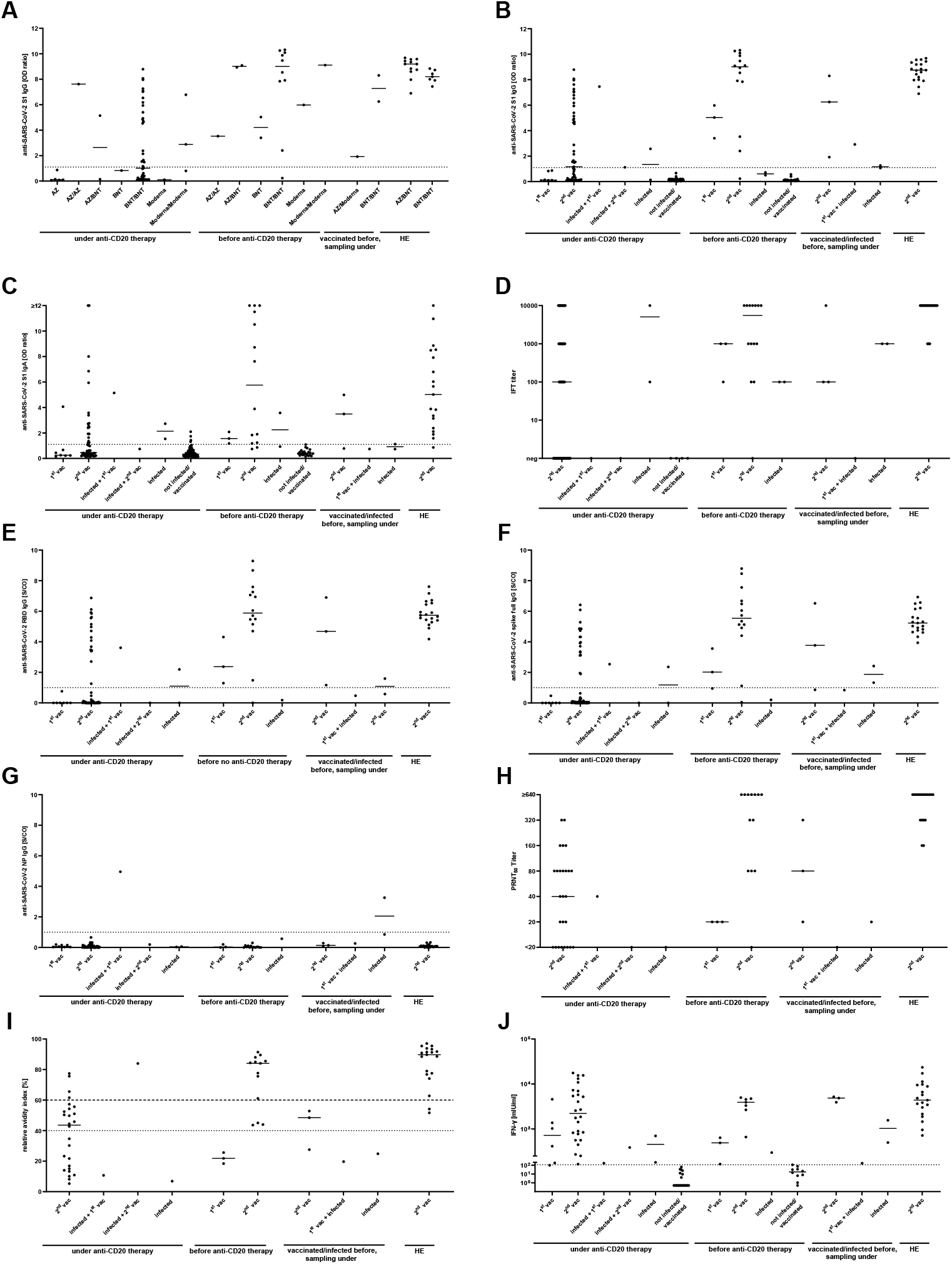
Details of humoral and cellular SARS-CoV-2 specific immune responses in patients with MS (pwMS) and healthy controls. (A) SARS-CoV-2 specific S1 IgG antibody responses after one or two vaccinations with different SARS-CoV-2 vaccines in anti-CD20 treated patients with MS (pwMS), pwMS before anti-CD20 therapy, pwMS vaccinated or infected before anti-CD20 therapy but sampled after start of anti-CD20 therapy, and HE. (B) Anti-SARS-CoV-2 S1 IgG, (C) Anti-SARS-CoV-2 S1 IgA, (D) IFT titer, (E) anti-RBD IgG, (F) anti-full spike IgG, (G) anti-NP IgG, (H) PRNT50 titer, (I) relative avidity index and (J) interferon-γ release in anti-CD20 treated pwMS, pwMS before anti-CD20 therapy, pwMS vaccinated or infected before anti-CD20 therapy but sampled after the start of anti-CD20 therapy, and HE. Dotted horizontal lines depict the manufacturer’s threshold for elevated anti-SARS-CoV-2 S1 IgG and anti-SARS-CoV-2 S1 IgA OD ratios (A-C), anti-SARS-CoV-2 RBD, full spike and NP IgG (E-G) and the tresholds of 40–60% for borderline and >60% for high antibody avidity (I). For IGRA, an arbitrary cutoff for positivity (dotted line) was set as a value of interferon-γ higher than the highest interferon-γ value in the not vaccinated/infected control group (111.41 mIU/ml) (J). Horizontal lines indicate the median. AZ = ChAdOx1 vaccine (Astrazeneca), BNT = BNT162b2 vaccine (Biontec), HE = hospital employees, IFN-γ = interferon-γ, IFT = immunofluorescence test, IgA = immunoglobulin A, IgG = immunoglobulin G, IGRA = interferon-γ release assay, IU = international units, OD = optical density, Moderna = mRNA-1273 vaccine, NP = nucleocapsid protein, ns = not significant, PRNT= plaque reduction neutralization test, RBD = receptor binding domain, S1 = SARS-CoV-2 spike protein S1 domain, S/Co= Signal to cut-off, vac = vaccination

We next analysed the functionality of SARS-CoV-2 specific antibodies. Only 18/26 (69.2%, 95%CI: 50.0-83.5%) anti-CD20 treated pwMS with reactive anti-SARS-CoV-2 S1 IgG as detected by ELISA exhibited neutralizing antibodies against an authentic SARS-CoV-2 isolate with a titer of 20 or greater compared to 12/13 (92.3%, 95%CI: 66.7-99.6%, *p*=0.23) pwMS before anti-CD20 therapy and 19/19 (100%, 95%CI: 83.2-100%, *p*=0.01) HE (Table 2). Accordingly, the neutralizing capacity of SARS-CoV-2 antibodies was lower in anti-CD20 treated pwMS (median [IQR] PRNT50 Titer: 40 [0-80]) than in pwMS before anti-CD20 therapy (PRNT50 Titer: 640 [80-640], *p*=0.006) and in HE (PRNT50 Titer: 640 [320-640], *p*<0.0001) (Fig 3B, Fig 4H). No differences in neutralizing capacity were found in pwMS before anti-CD20 therapy and HE (*p*=0.74).

Impaired functionality of SARS-CoV-2 antibodies was also reflected in the maturation of IgG avidity: After two vaccinations, only 4/26 (15.4%, 95%CI: 6.2-33.5%) anti-CD20 treated pwMS had high anti-SARS-CoV-2 S1 IgG avidity indices compared to 10/13 (76.9%, 95%CI: 49.7-91.8%, *p*=0.0003) pwMS before anti-CD20 therapy and 17/19 (85.5%, 95%CI: 68.6-98.1%, *p*<0.0001) HE (Table 2). Accordingly, the median (IQR) relative avidity index was lower in anti-CD20 treated pwMS (43.6% [14.8-54.6%]) than in pwMS before anti-CD20 therapy (84.1% [53.1-86.8%], *p*=0.0006) and in HE (89.7 [76.8-93.4%], *p*=0.003, Fig 3C, Fig 4I). No differences in maturation of IgG avidity were found in pwMS before anti-CD20 therapy and HE (*p*=0.36).

### Cellular immune response to SARS-CoV-2 vaccinations

Cellular immune responses following two SARS-CoV-2 vaccinations were studied by an SARS-CoV-2 spike protein specific IGRA in all pwMS with a lithium heparinized blood sample available and in all HE. Remarkably, SARS-CoV-2 spike protein specific T cell responses were detectable in all anti-CD20 treated pwMS (26/26, 100%, 95%CI: 87.1-100%), similar to pwMS before anti-CD20 therapy (7/7, 100%, 95%CI: 64.6-100%, *p*=1), and HE (19/19, 100%, 95%CI: 83.2-100%, *p*=1, Table 2). Accordingly, levels of IFN-γ released by SARS-CoV-2 spike protein specific T cells were similar between the three groups (Fig 3D, Fig 4J). Levels of SARS-CoV-2 spike protein specific T cell responses did not correlate with the level of anti-SARS-CoV-2 S1 IgG antibodies (*r*=-0.21, *p*=0.31) or the interval between last anti-CD20 treatment and the second SARS-CoV-2 vaccination (*r*=-0.31, *p*=0.13).

Altogether, following two vaccinations against SARS-CoV-2, across all humoral immune response parameters, lower proportions of positive outcomes were detected in anti-CD20 treated pwMS than in pwMS before anti-CD20 therapy or in HE (Fig 3E, Table 2). In contrast, SARS-CoV-2 specific T cells were detected in all twice-vaccinated anti-CD20 treated pwMS, indicating the generation of a robust cellular immune response.

### Parameters associated with vaccine-induced antibody responses in anti-CD20 treated pwMS

When analysing all 51 twice-vaccinated anti-CD20 treated pwMS, levels of anti-SARS-CoV-2 S1 IgG increased with increasing time between the last anti-CD20 therapy and the second vaccination (*r*=0.42, *p*=0.002, Fig 5A). Of note, after an interval of >279 days between the last anti-CD20 therapy and the second vaccination, no anti-SARS-CoV-2 S1 IgG negative patients were detected. Furthermore, in the 26 anti-SARS-CoV-2 S1 IgG antibody reactive anti-CD20 treated pwMS vaccinated twice, a strong positive correlation was observed between the relative avidity indices and the interval between the last anti-CD20 therapy and the second vaccination (*r*=0.70, *p*<0.001, Fig 5B). Likewise, there was a positive correlation between the PRNT50 titer and the interval between the last anti-CD20 therapy and the second vaccination (*r*=0.44, *p*=0.03, Fig 5C).

**Figure 5.**
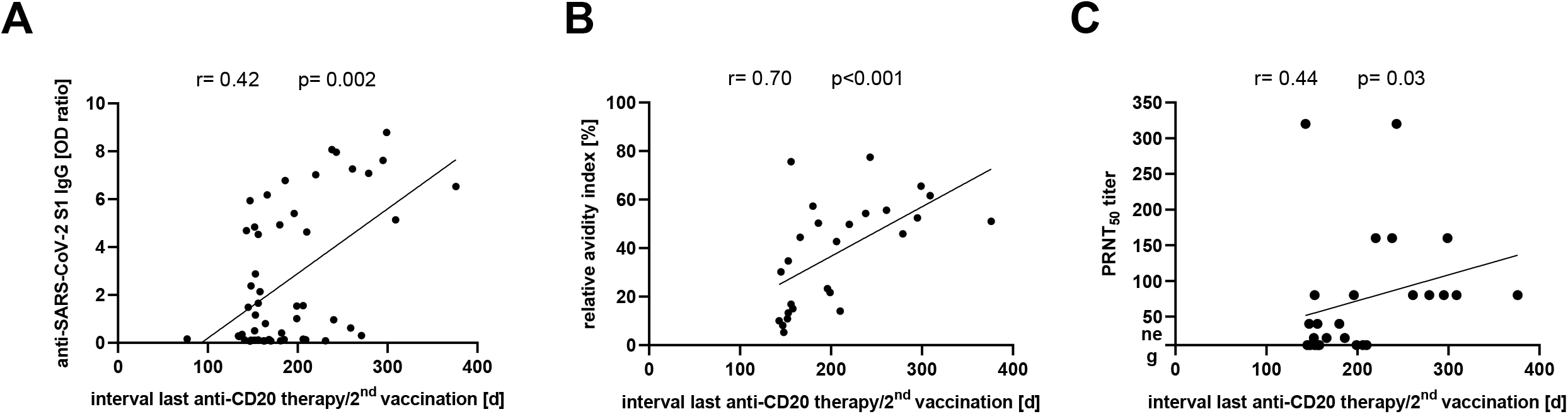
(A) Correlation of anti-SARS-CoV-2 S1 IgG OD ratio and interval from the last anti-CD20 therapy to the second SARS-CoV-2 vaccination in anti-CD20 treated pwMS (n=51). (B) Correlation of relative avidity index and (C) PRNT50 titer with the interval from the last anti-CD20 therapy to the second SARS-CoV-2 vaccination in anti-SARS-CoV-2 S1 IgG reactive anti-CD20 treated pwMS (n=26). Correlations were calculated by Spearman’s method. d = days, IgG = immunoglobulin G, OD = optical density, PRNT = plaque reduction neutralization test, S1 = SARS-CoV-2 spike protein S1 domain

In contrast, anti-SARS-CoV-2 S1 IgG levels were not associated with age (*r*=-0.007, *p*=0.96) or time from the second vaccination to blood withdrawal (*r*=-0.05, *p*=0.75). Likewise, there was no association of relative avidity indices (*r*=0.18, *p*=0.46) or the PRNT50 titer (*r*=0.15, *p*=0.54) with the interval from the second vaccination to blood withdrawal. Furthermore, no associations were observed between anti-SARS-CoV-2 S1 IgG antibody levels and total serum IgG levels (*r*=0.18, *p*=0.25) or the cumulative lifetime dose of anti-CD20 therapy (*r*=0.04, *p*=0.78).

### SARS-CoV-2 infections in patients with multiple sclerosis

Infection with SARS-CoV-2 was defined as detection of reactive SARS-CoV-2 antibodies in at least two of four antibody test systems (anti-SARS-CoV-2 S1 IgG ELISA, anti-SARS-CoV-2 S1 IgA ELISA, spike protein IFT, and SeraSpot® anti-SARS-CoV-2 IgG) or a positive IGRA in pwMS not vaccinated against SARS-CoV-2. According to this definition, nine pwMS were identified as infected with SARS-CoV-2. Five of these nine pwMS (two women, three men; age range 24-68 years) were infected with SARS-CoV-2 before initiation of anti-CD20 therapy. In these five patients, symptoms of SARS-CoV-2 infections were overall mild to moderate and none of the patients needed hospitalization. Of note, three of these five patients developed the first clinical symptoms of MS between 2-6 months after symptoms of SARS-CoV-2 infection. Four of nine SARS-CoV-2 infected pwMS (one woman, three men; age range 18-51 years) were treated with anti-CD20 therapies at the time they had symptoms compatible with COVID-19. While in three of these patients symptoms of COVID-19 were mild to moderate, one patient had a more severe course, including worsening of pre-existing MS symptoms, requiring hospitalization but no intensive care treatment. Clinical details and results of SARS-CoV-2 antibody and T cell assessments of all nine SARS-CoV-2 infected pwMS are summarized in Table 3. Altogether, all 9 patients recovered fully and none of the 9 SARS-CoV-2 infected patients died of COVID-19. Antibodies against the SARS-CoV-2 nucleocapsid, indicating past SARS-CoV-2 infection, were detected in only two SARS-CoV-2 infected pwMS, one was infected before anti-CD20 therapy and one under anti-CD20 therapy. Importantly, in the two anti-CD20 treated SARS-CoV-2 infected pwMS in whom SARS-CoV-2 specific T cell responses could be measured, SARS-CoV-2 specific T cells were detectable.

**Table 3.**
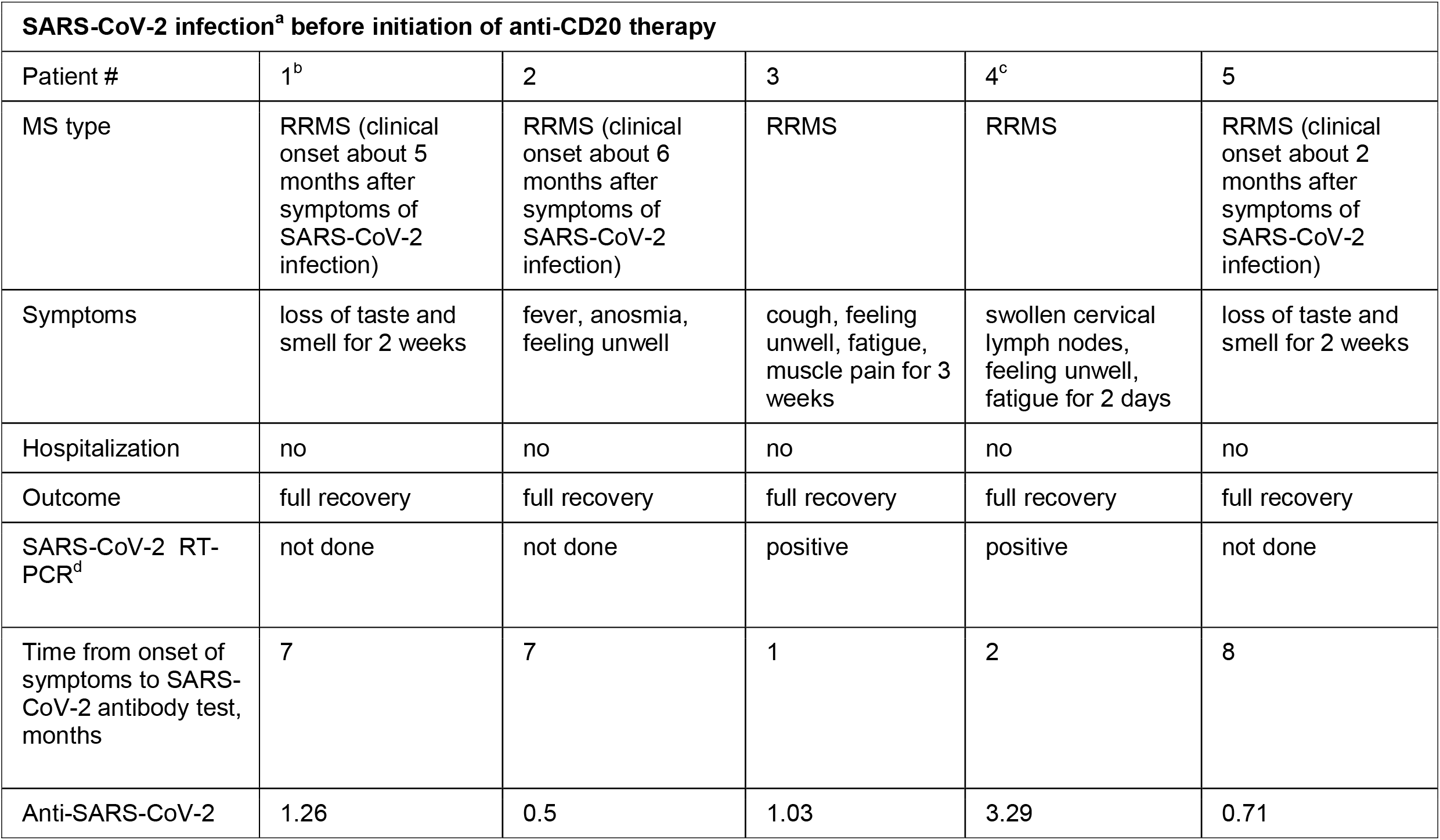
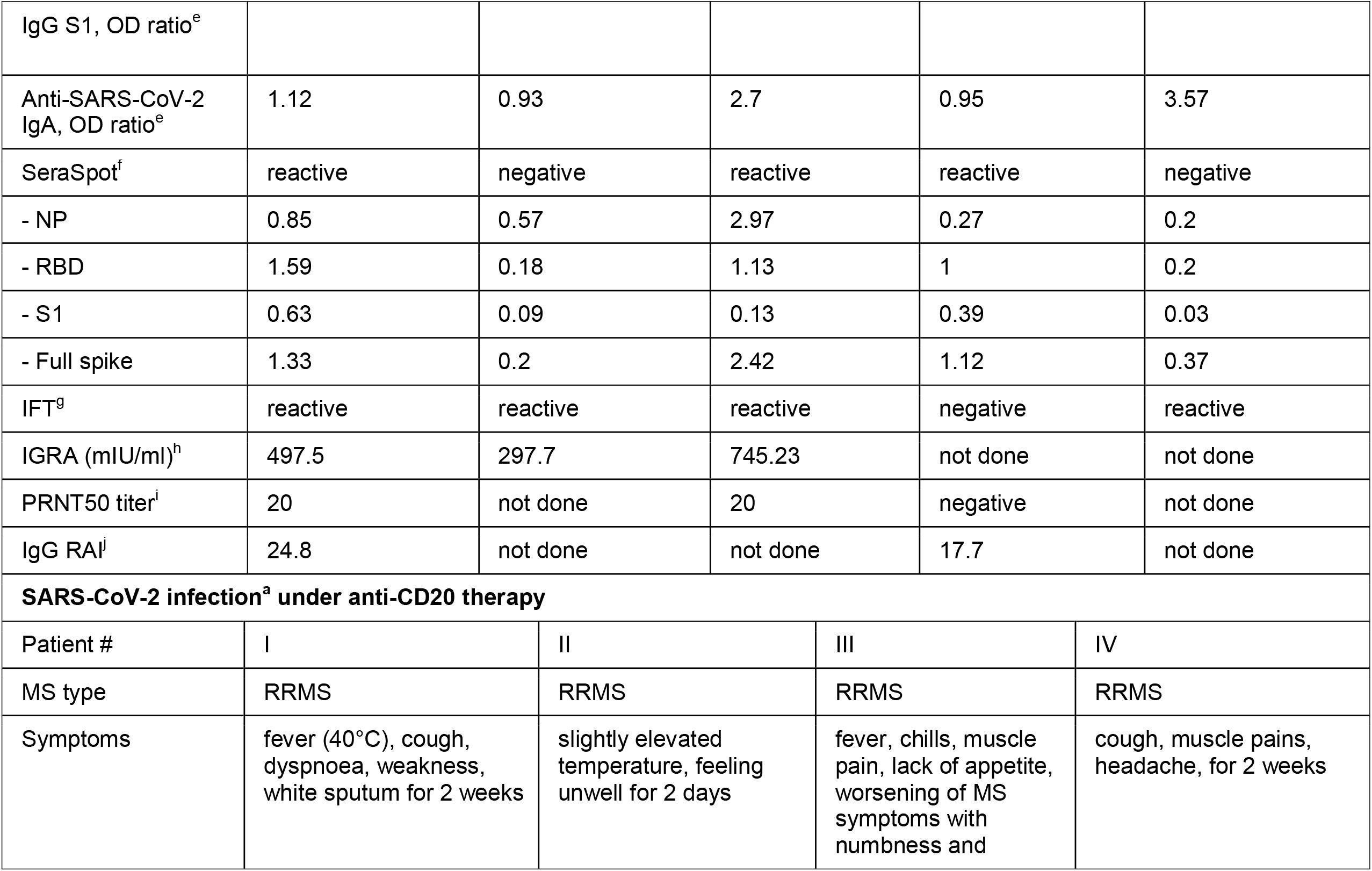
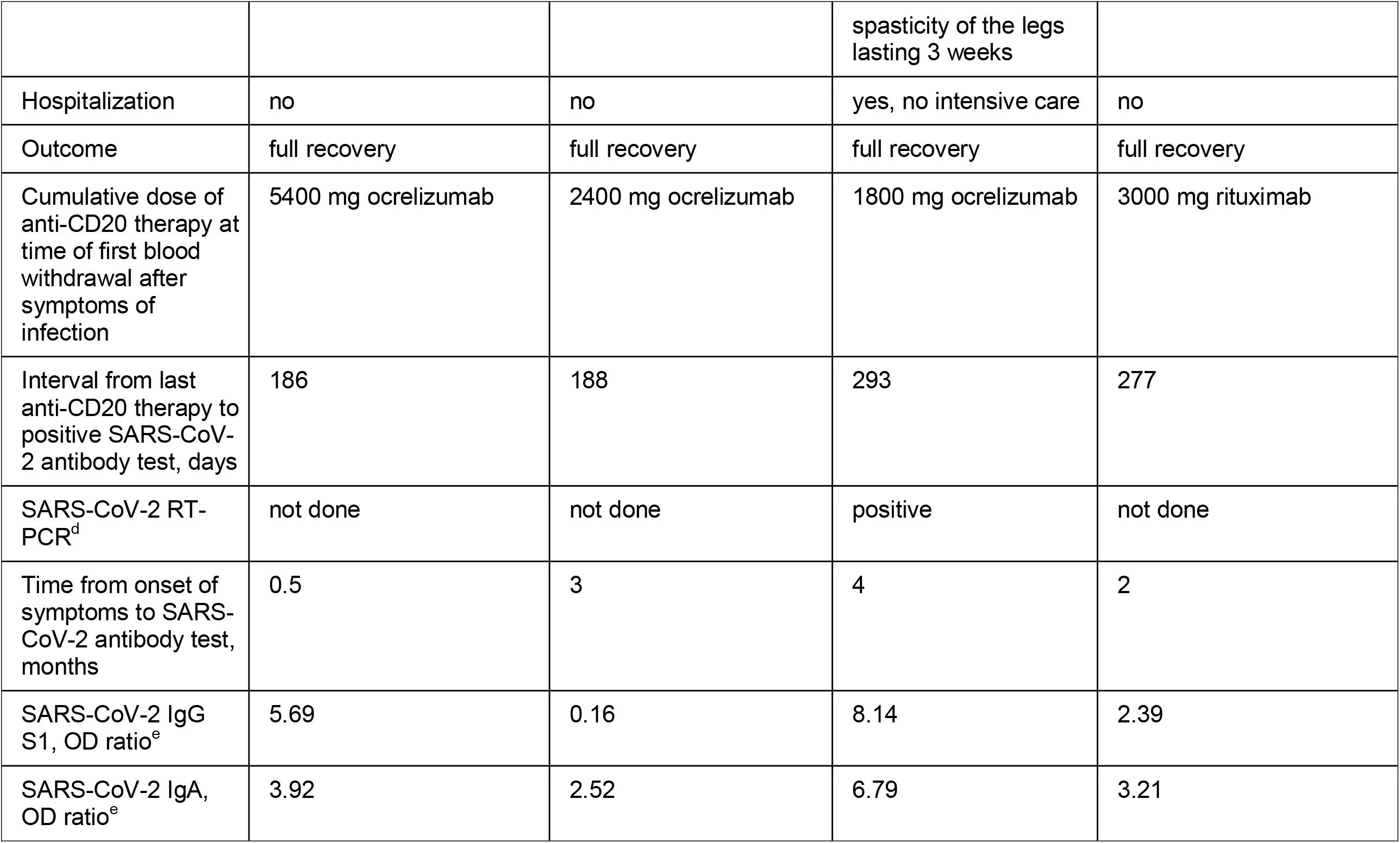
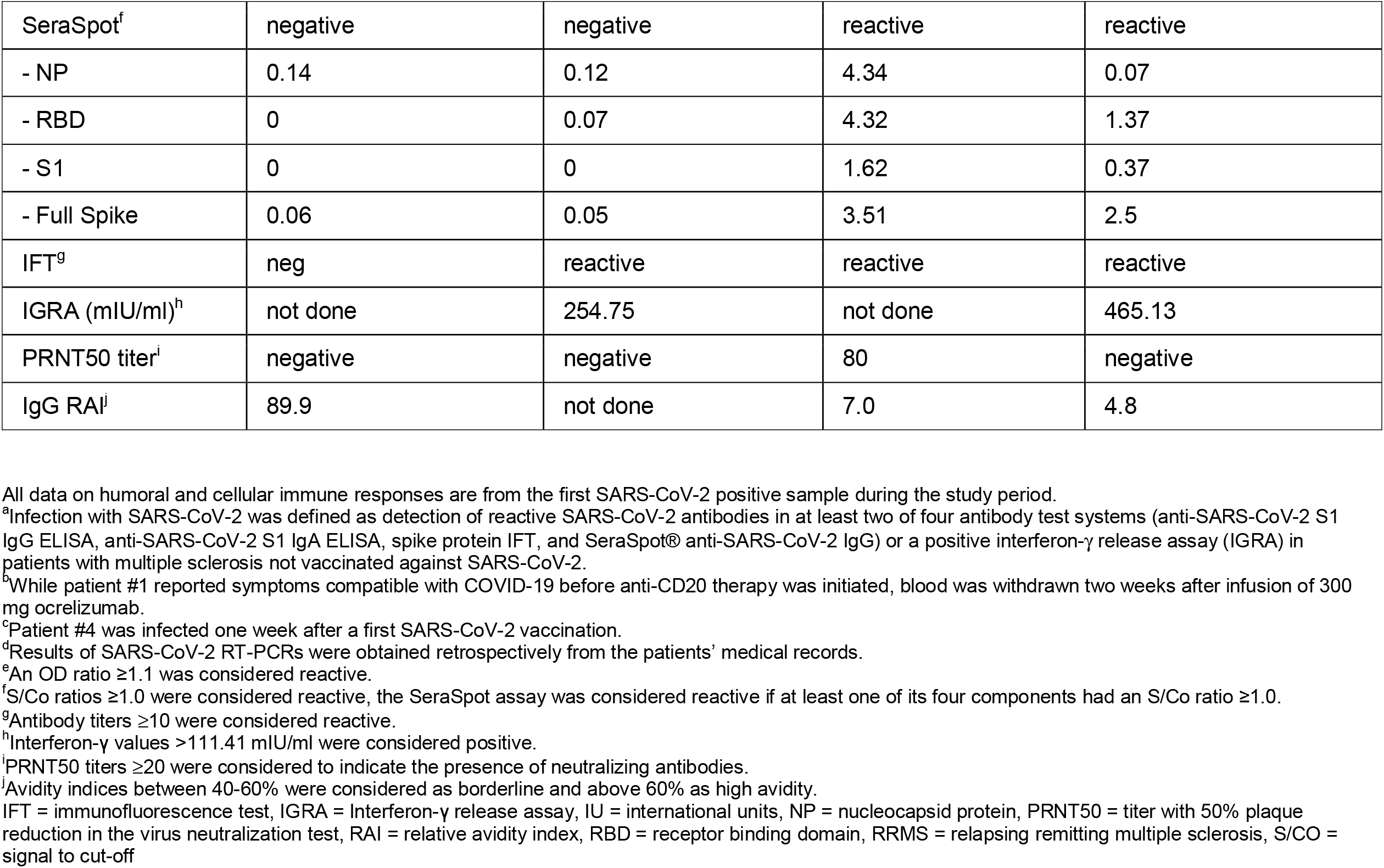
Clinical, treatment and serological characteristics of SARS-CoV-2 infected patients with multiple sclerosis.

| **SARS-CoV-2 infection<sup>a</sup> before initiation of anti-CD20 therapy** |  |  |  |  |  |
| --- | --- | --- | --- | --- | --- |
| Patient # | 1<sup>b</sup> | 2 | 3 | 4<sup>c</sup> | 5 |
| MS type | RRMS (clinical onset about 5 months after symptoms of SARS-CoV-2 infection) | RRMS (clinical onset about 6 months after symptoms of SARS-CoV-2 infection) | RRMS | RRMS | RRMS (clinical onset about 2 months after symptoms of SARS-CoV-2 infection) |
| Symptoms | loss of taste and smell for 2 weeks | fever, anosmia, feeling unwell | cough, feeling unwell, fatigue, muscle pain for 3 weeks | swollen cervical lymph nodes, feeling unwell, fatigue for 2 days | loss of taste and smell for 2 weeks |
| Hospitalization | no | no | no | no | no |
| Outcome | full recovery | full recovery | full recovery | full recovery | full recovery |
| SARS-CoV-2 RT-PCR<sup>d</sup> | not done | not done | positive | positive | not done |
| Time from onset of symptoms to SARS-CoV-2 antibody test, months | 7 | 7 | 1 | 2 | 8 |
| Anti-SARS-CoV-2 | 1.26 | 0.5 | 1.03 | 3.29 | 0.71 |
| IgG S1, OD ratio<sup>e</sup> |  |  |  |  |  |
| Anti-SARS-CoV-2 IgA, OD ratio<sup>e</sup> | 1.12 | 0.93 | 2.7 | 0.95 | 3.57 |
| SeraSpot<sup>f</sup> | reactive | negative | reactive | reactive | negative |
| - NP | 0.85 | 0.57 | 2.97 | 0.27 | 0.2 |
| - RBD | 1.59 | 0.18 | 1.13 | 1 | 0.2 |
| - S1 | 0.63 | 0.09 | 0.13 | 0.39 | 0.03 |
| - Full spike | 1.33 | 0.2 | 2.42 | 1.12 | 0.37 |
| IFT<sup>g</sup> | reactive | reactive | reactive | negative | reactive |
| IGRA (mIU/ml)<sup>h</sup> | 497.5 | 297.7 | 745.23 | not done | not done |
| PRNT50 titer<sup>i</sup> | 20 | not done | 20 | negative | not done |
| IgG RAI<sup>j</sup> | 24.8 | not done | not done | 17.7 | not done |
| **SARS-CoV-2 infection<sup>a</sup> under anti-CD20 therapy** |  |  |  |  |  |
| Patient # | I | II | III | IV |  |
| MS type | RRMS | RRMS | RRMS | RRMS |  |
| Symptoms | fever (40°C), cough, dyspnoea, weakness, white sputum for 2 weeks | slightly elevated temperature, feeling unwell for 2 days | fever, chills, muscle pain, lack of appetite, worsening of MS symptoms with numbness and | cough, muscle pains, headache, for 2 weeks |  |

|  |  |  |  |  |
| --- | --- | --- | --- | --- |
|  |  |  | spasticity of the legs<br>lasting 3 weeks |  |
| Hospitalization | no | no | yes, no intensive care | no |
| Outcome | full recovery | full recovery | full recovery | full recovery |
| Cumulative dose of<br>anti-CD20 therapy at<br>time of first blood<br>withdrawal after<br>symptoms of<br>infection | 5400 mg ocrelizumab | 2400 mg ocrelizumab | 1800 mg ocrelizumab | 3000 mg rituximab |
| Interval from last<br>anti-CD20 therapy to<br>positive SARS-CoV-2 antibody test, days | 186 | 188 | 293 | 277 |
| SARS-CoV-2 RT-PCR<sup>d</sup> | not done | not done | positive | not done |
| Time from onset of<br>symptoms to SARS-CoV-2 antibody test,<br>months | 0.5 | 3 | 4 | 2 |
| SARS-CoV-2 IgG<br>S1, OD ratio<sup>e</sup> | 5.69 | 0.16 | 8.14 | 2.39 |
| SARS-CoV-2 IgA,<br>OD ratio<sup>e</sup> | 3.92 | 2.52 | 6.79 | 3.21 |
| SeraSpot<sup>f</sup> | negative | negative | reactive | reactive |
| - NP | 0.14 | 0.12 | 4.34 | 0.07 |
| - RBD | 0 | 0.07 | 4.32 | 1.37 |
| - S1 | 0 | 0 | 1.62 | 0.37 |
| - Full Spike | 0.06 | 0.05 | 3.51 | 2.5 |
| IFT<sup>g</sup> | neg | reactive | reactive | reactive |
| IGRA (mIU/ml)<sup>h</sup> | not done | 254.75 | not done | 465.13 |
| PRNT50 titer<sup>i</sup> | negative | negative | 80 | negative |
| IgG RAI<sup>j</sup> | 89.9 | not done | 7.0 | 4.8 |

All data on humoral and cellular immune responses are from the first SARS-CoV-2 positive sample during the study period.
<sup>a</sup>Infection with SARS-CoV-2 was defined as detection of reactive SARS-CoV-2 antibodies in at least two of four antibody test systems (anti-SARS-CoV-2 S1 IgG ELISA, anti-SARS-CoV-2 S1 IgA ELISA, spike protein IFT, and SeraSpot® anti-SARS-CoV-2 IgG) or a positive interferon-γ release assay (IGRA) in patients with multiple sclerosis not vaccinated against SARS-CoV-2.
<sup>b</sup>While patient #1 reported symptoms compatible with COVID-19 before anti-CD20 therapy was initiated, blood was withdrawn two weeks after infusion of 300 mg ocrelizumab.
<sup>c</sup>Patient #4 was infected one week after a first SARS-CoV-2 vaccination.
<sup>d</sup>Results of SARS-CoV-2 RT-PCRs were obtained retrospectively from the patients' medical records.
<sup>e</sup>An OD ratio ≥1.1 was considered reactive.
<sup>f</sup>S/Co ratios ≥1.0 were considered reactive, the SeraSpot assay was considered reactive if at least one of its four components had an S/Co ratio ≥1.0.
<sup>g</sup>Antibody titers ≥10 were considered reactive.
<sup>h</sup>Interferon-γ values >111.41 mIU/ml were considered positive.

## Discussion

The key findings of this comprehensive analysis of SARS-CoV-2 specific humoral and cellular immune responses in a large cohort of pwMS monitored throughout the first 17 months of the SARS-CoV-2 pandemic are (1) that levels as well as functionality of antibody responses to SARS-CoV-2 vaccinations were diminished in anti-CD20 treated pwMS, (2) that this effect was attenuated with increasing time from the last anti-CD20 infusion to the second vaccination and (3) that anti-CD20 treated pwMS developed robust SARS-CoV-2 specific T cell responses following SARS-CoV-2 vaccinations and infections.

The present results extend results of previous studies, which showed diminished anti-SARS-CoV-2 antibody levels following SARS-CoV-2 vaccinations in anti-CD20 treated pwMS,^5,6,9^ by demonstrating that not only antibody levels, but also the functionality of SARS-CoV-2 antibodies is reduced in anti-CD20 treated pwMS. The capacity of SARS-CoV-2 antibodies to neutralize SARS-CoV-2 was lower in twice-vaccinated anti-CD20 treated pwMS as compared to twice-vaccinated pwMS before initiation of anti-CD20 therapies and HE, consistent with recent findings in anti-CD20 treated pwMS obtained with a different SARS-CoV-2 neutralization assay.^10^ As levels of neutralizing antibodies to SARS-CoV-2 were shown to predict protection from symptomatic SARS-CoV-2 infections^15^, these results suggest that humoral immune protection against SARS-CoV-2 may be reduced in anti-CD20 treated pwMS. Furthermore, the diminished avidity of SARS-CoV-2 antibodies in anti-CD20 treated pwMS indicates that B cell depletion by anti-CD20 therapies interferes with the processes of normal antibody maturation. While the mechanisms underlying impaired antibody maturation in B cell depleted pwMS remain to be clarified, both, the reduced neutralization capacity and the diminished antibody avidity, might contribute to an attenuated humoral immune response against SARS-CoV-2, but also against other vaccinations in anti-CD20 treated pwMS.^4^

Importantly, SARS-CoV-2 antibody levels, avidity and neutralizing capacity increased with increasing time from the last anti-CD20 therapy to the second SARS-CoV-2 vaccination. This is most likely explained by increasing numbers of reappearing B cells with increasing time after anti-CD20 infusions. Indeed, the interval from the last anti-CD20 therapy to the second SARS-CoV-2 vaccination of >279 days after which no negative SARS-CoV-2 IgG anti-CD20 treated pwMS were identified is consistent with slow B cell repopulation starting about 6 months after the last anti-CD20 infusion.^23^

From a clinical perspective, these findings suggest that to enhance humoral immune responses, SARS-CoV-2 vaccinations should be applied as late as possible within the 6 monthly infusion cycles of intravenous anti-CD20 therapy. Furthermore, if clinically justified, one may consider prolonging the infusion interval between anti-CD20 infusions to increase chances of successful SARS-CoV-2 vaccinations.^24^ Finally, the reduced SARS-CoV-2 antibody responses in anti-CD20 treated pwMS suggest that an additional booster vaccination may be considered in this patient population.

Remarkably, in all anti-CD20 treated pwMS vaccinated twice and in all SARS-CoV-2 infected pwMS in whom T cell responses could be measured, SARS-CoV-2 specific T cells were detectable. Together with very recent similar results^9,10^, these data suggest that B cell depletion in anti-CD20 treated pwMS does not impair the generation of SARS-CoV-2 specific cellular immune responses. As an early and robust SARS-CoV-2 specific T cell response is associated with mild or asymptomatic SARS-CoV-2 infections even in the absence of antibodies ^25,26^, these findings indicate that SARS-CoV-2 vaccinations of anti-CD20 treated pwMS may result in some degree of protection from COVID-19. Together with data showing that SARS-CoV-2 variants of concern partially escaping humoral immunity can still be recognized by SARS-CoV-2 specific T cells^27^, these findings clearly support current recommendations to vaccinate anti-CD20 treated pwMS against COVID-19.^2^ Generation of SARS-CoV-2 specific T cells might also have contributed to recovery from SARS-CoV-2 infections in previously reported anti-CD20 treated pwMS infected with SARS-CoV-2, who did not develop anti-SARS-CoV-2 antibodies.^7,8,28^ Nevertheless, further studies on the importance of T cells for protection from severe COVID-19 and their role in clearance of SARS-CoV-2 in anti-CD20 treated pwMS are needed. Beyond SARS-CoV-2, preserved T cell response after mRNA and adenovirus-based vaccines suggest a possible advantage of these vaccine types in patients under B cell depleting therapies and may give reason to further investigate these vaccine types for other pathogens in anti-CD20 treated patients.

Protective immunity against SARS-CoV-2 depends on characteristics of circulating virus strains and the build-up of antibody levels, specific T cells, and immune memory.^13,14,29^ Easily assessable correlates of protection, such as SARS-CoV-2 specific antibody or T cell cut-off values, have therefore so far not been and will possibly not be identified. To analyse the clinical significance of reduced antibody levels, but preserved T cell responses against SARS-CoV-2, longitudinal studies of SARS-CoV-2 breakthrough infections in anti-CD20 treated pwMS vaccinated against SARS-CoV-2 will be required.^30^

Based on the detection of SARS-CoV-2 antibodies, we identified four anti-CD20 treated pwMS with SARS-CoV-2 infections. However, given that some anti-CD20 treated pwMS with RT-PCR confirmed SARS-CoV-2 infections may not develop anti-SARS-CoV-2 antibodies^7,8,28^, the number of SARS-CoV-2 infected anti-CD20 treated pwMS identified in our cohort could be an underestimate. Interestingly, the detection rate of SARS-CoV-2 infections was lower in anti-CD20 treated pwMS (4/175, 2.3%) than in pwMS before initiation of anti-CD20 therapy (5/47, 10.6%), which could be related to less frequent antibody responses in anti-CD20 treated pwMS. However, an additional explanation for this finding might be more cautious behavior of pwMS receiving anti-CD20 therapy, such as stricter mask wearing and avoidance of potentially risky situations. Remarkably, in three of five patients infected with SARS-CoV-2 before anti-CD20 therapy, first symptoms of MS develeoped few months after symptoms of SARS-CoV-2 infections, similar to previously described cases.^31^ The relatively frequent detection of SARS-CoV-2 infections in pwMS before anti-CD20 therapy in our cohort could therefore also be due to potential triggering of the first clinical manifestation of MS by SARS-CoV-2 infections in these three patients.^32^ Importantly, in both anti-CD20 treated SARS-CoV-2 infected pwMS in whom SARS-CoV-2 specific T cells could be determined, SARS-CoV-2 specific T cells were present, indicating that, similar to SARS-CoV-2 vaccinated anti-CD20 treated pwMS, SARS-CoV-2 infected anti-CD20 treated pwMS can generate SARS-CoV-2 specific T cells.

Limitations of this observational study include lack of data on total lymphocyte and B cell counts at the time of SARS-CoV-2 vaccinations precluding evaluations of these parameters as potential risk factors for low vaccine immunogenicity. Furthermore, given the limited follow-up time of the present study and potentially decreasing SARS-CoV-2 immune responses over time, it will be important to analyse anti-SARS-CoV-2 antibody and T cell responses in anti-CD20 treated patients also in the long term. Finally, the low numbers of pwMS vaccinated with vaccines other than BNT162b2 precluded formal comparisons of the immunogenicity of different vaccines.

Altogether, this study shows that although levels and functionality of anti-SARS-CoV-2 antibodies are diminished, SARS-CoV-2 specific T cell responses are preserved in SARS-CoV-2 vaccinated or infected anti-CD20 treated pwMS. Preserved T cell responses suggest that anti-CD20 treated pwMS develop at least some degree of protection from COVID-19, supporting SARS-CoV-2 vaccinations in anti-CD20 treated pwMS. A longer interval between anti-CD20 infusions and vaccination may enhance the extent and the functionality of SARS-CoV-2 antibody responses. These findings should inform treatment decisions and management of SARS-CoV-2 vaccinations in pwMS.

## Data Availability

All data produced in the present study are contained in the manuscrip or are available upon reasonable request to the authors.

## Acknowledgements

We thank Betina Jaenicke, Anita Kaiser-Friedrich, Marie Luisa Schmidt, Patricia Tscheak, Julia Tesch, Johanna Riege, Petra Mackeldanz, and Felix Walper for excellent assistance. Parts of this work were supported by grants from the Berlin Institute of Health (BIH) and Berlin University Alliance to CD and VMC. This study was further supported by the German Ministry of Education and Research through Forschungsnetzwerk der Universitätsmedizin zu COVID-19, COVIM, FKZ: 01KX2021 to CD and VMC, and projects VARIPath (01KI2021) to VMC. VMC is a participant in the BIH–Charité Clinician Scientist Program funded by Charité -Universitätsmedizin Berlin and the Berlin Institute of Health. KR is a participant in the BIH Clinical Fellow Program funded by Stiftung Charité. FP is a participant in the BIH-Charité Clinician Scientist Program funded by Charité -Universitätsmedizin Berlin and the Berlin Institute of Health.

## Authors Contributions

Conception and design of the study: V.M.C., K.R.

Acquisition and analysis of data: T.S., C.O., F.P., P.S., M.N., F.S., C.D., V.M.C., K.R.

Drafting of the manuscript or figures: T.S., C.O., V.M.C., K.R.

